# Sequential epiretinal stimulation improves discrimination in simple shape discrimination tasks only

**DOI:** 10.1101/2022.03.08.22270800

**Authors:** Breanne Christie, Roksana Sadeghi, Arathy Kartha, Avi Caspi, Francesco V. Tenore, Roberta L. Klatzky, Gislin Dagnelie, Seth Billings

## Abstract

**Objective:** Electrical stimulation of the retina can elicit flashes of light called phosphenes, which can be used to restore rudimentary vision for people with blindness. Functional sight requires stimulation of multiple electrodes to create patterned vision, but phosphenes tend to merge together in an uninterpretable way. Sequentially stimulating electrodes in human visual cortex has recently demonstrated that shapes could be “drawn” with better perceptual resolution relative to simultaneous stimulation. The goal of this study was to evaluate if sequential stimulation would also form clearer shapes when the retina is the neural target.

**Approach:** Two human participants with retinitis pigmentosa who had Argus® II retinal prostheses participated in this study. We evaluated different temporal parameters for sequential stimulation in phosphene shape mapping and forced-choice discrimination tasks. For the discrimination tasks, performance was compared between stimulating electrodes simultaneously versus sequentially.

**Main results:** Phosphenes elicited by different electrodes were reported as vastly different shapes. Sequential electrode stimulation outperformed simultaneous stimulation in simple discrimination tasks, in which shapes were created by stimulating 3-4 electrodes, but not in more complex discrimination tasks involving 5+ electrodes. For sequential stimulation, the optimal pulse train duration was 200 ms when stimulating at 20 Hz and the optimal gap interval was tied between 0 and 50 ms. Efficacy of sequential stimulation also depended strongly on selecting electrodes that elicited phosphenes with similar shapes and sizes.

**Significance:** An epiretinal prosthesis can produce coherent simple shapes with a sequential stimulation paradigm, which can be used as rudimentary visual feedback. However, success in creating more complex shapes, such as letters of the alphabet, is still limited. Sequential stimulation may be most beneficial for epiretinal prostheses in simple tasks, such as basic navigation, rather than complex tasks such as object identification.

## Introduction

Different neural targets along the visual pathway can be stimulated to elicit flashes of light known as phosphenes, which can be used as rudimentary visual feedback for people with blindness. To date, visual neuroprostheses have been implanted for at least several months in the retina (Humayun et al. 2012; Stingl et al. 2013; Fujikado et al. 2016; Petoe et al. 2021; Barnes et al. 2016; Palanker et al. 2022; Xu et al. 2021), optic nerve (Veraart et al. 2003), and visual cortex of human subjects (Beauchamp et al. 2020; Fernández et al. 2021). The Argus® II (Second Sight Medical Products, Sylmar, CA, USA) is a device that was implanted commercially in the United States from 2014 through 2020. It features a 6×10 electrode array implanted on the epiretinal surface. Visual information regarding an Argus user’s surroundings is provided by a small camera mounted to a pair of glasses and a body-worn video processor (Figure 1a). When receiving visual input from an Argus device, people with blindness improved on orientation and mobility tasks based on walking towards a high-contrast target and walking along a high-contrast line on the floor (Humayun et al. 2012). Their performance also improved for object localization and motion discrimination tasks (Ahuja et al. 2011; Barry, Dagnelie, and Argus II Study Group 2012; Dorn et al. 2013; Dagnelie et al. 2017).

**Figure 1:**
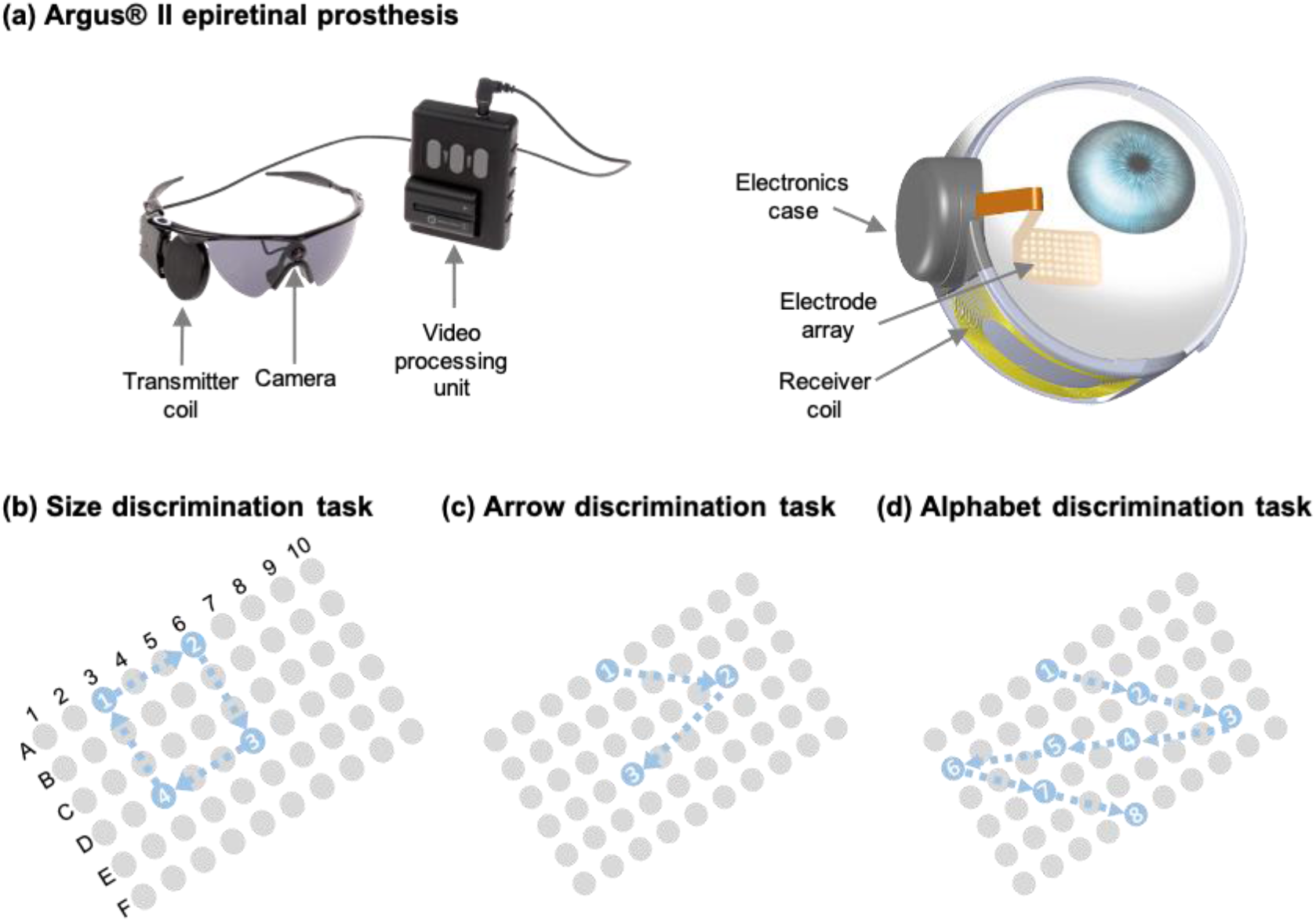
Experimental methods. **(a)** Two people with retinitis pigmentosa were implanted with Argus® II epiretinal prostheses. The external components (left) consist of a video processing unit and glasses that contain a transmitter coil and camera. The internal components (right) consist of an electronics case, receiver coil, and electrode array planted on the epiretinal surface. Though not pictured, the array is implanted at a ∼45° angle with respect to the horizontal meridian. Images are courtesy of Second Sight Medical Products, Inc. **(b)** Both participants performed a size discrimination task, in which they were presented with two squares and had to choose which square was larger. Performance was compared when participants used simultaneous stimulation vs. sequential stimulation. In this example, blue circles represent the four stimulated electrodes that produce a square with each corner separated by two electrodes. The gray circles represent inactive electrodes. The white numbers within active electrode circles represent the order of presentation under the sequential stimulation paradigm. **(c)** The participants performed an arrow discrimination task, in which they were presented with one arrow and had to choose if it was pointing to the left or right. Performance was compared when participants used simultaneous stimulation vs. sequential stimulation. In this example, the blue circles represent three stimulated electrodes that created a right-pointing arrow. **(d)** Both participants performed an alphabet letter discrimination task, in which they were presented with a letter and had to choose if it was the letter O, V, K, or Z. Blue circles represent the electrodes used to produce the letter Z, for example. Performance was compared when participants used simultaneous stimulation vs. sequential stimulation.

The visual feedback provided by neural prostheses is still quite limited, however. The information delivered by a single phosphene is minimal, so patterned vision is used to maximize real-time visual understanding of one’s environment. Patterned vision is created by stimulating through multiple electrodes within the array. In theory, stimulating electrodes that form a specific shape within an array should elicit phosphenes in the same shape, given that the image projection on the retina is formed by geometric optics. However, it is not typically that simple: when multiple electrodes are stimulated simultaneously, phosphenes tend to merge together in complex ways (Bosking et al. 2018; Rizzo et al. 2003; Horsager et al. 2011; Horsager, Greenberg, and Fine 2010; Wilke et al. 2011). For example, Argus II devices enabled only ∼50% of test subjects to reliably read large-print letters (41° or ∼22.6 cm when viewed from 30 cm away), and single-letter recognition took anywhere between six seconds to 3.5 minutes (da Cruz et al. 2013). Moreover, the highest level of visual acuity achieved is only about 20/1260 (Humayun et al. 2012), which is dictated by the 550 µm electrode spacing. Some tasks can be performed by scanning the field-of-view of the camera (Caspi and Zivotofsky 2015; Hallum and Dakin 2021) or the region-of-interest using eye movements (Caspi et al. 2018). However, scanning limits the performance of the system (Peli 2020), making it critical to develop stimulation paradigms that better enable patterned sight.

To improve the clarity of images produced by stimulating through multiple electrodes, recent studies tested a stimulation paradigm that sequentially and rapidly stimulated individual electrodes placed on human visual cortex (Beauchamp et al. 2020; Oswalt et al. 2021). They found that sequential stimulation produced shapes that were recognized more quickly and accurately than traditional simultaneous stimulation of electrodes. To evaluate if a similar procedure would improve object recognition for retinal prosthesis users, we performed phosphene shape mapping experiments and forced choice discrimination tasks to evaluate if Argus II users could discriminate between differently sized squares, arrow direction, and letters of the alphabet. We hypothesized that sequential stimulation would outperform simultaneous stimulation in all discrimination tasks.

## Methods

### Human participants with Argus II retinal prostheses

The following experiments were conducted with two individuals (S1 and S2) with retinitis pigmentosa. Both participants had been implanted with unilateral Argus II retinal prostheses for over seven years. The arrays were centered over the fovea with electrodes oriented at an approximately 45° angle relative to the horizontal meridian (Gregori, Davis, and Rizzo 2016). The study protocol was approved by the Johns Hopkins Medicine Institutional Review Board. Informed consent was obtained prior to participation in research-related activities.

The detection threshold for each electrode and a single slope of the psychometric function were determined by a Bayesian adaptive method in a yes/no experiment using a Weibull function. In each trial, stimulation was followed by a sound cue and the responses were collected using two keys on a gaming remote. Based on the response in each trial, the prior probabilities were updated, and the next trial stimulus amplitude level was presented at the level that provided the most information about the threshold and slope of the Weibull function. The thresholds for each electrode and one slope were estimated concurrently for all electrodes within 30 trials per electrode (Kontsevich and Tyler 1999). In an attempt to approximately match phosphene brightness between electrodes and across trials, stimulation current for each electrode was set to a suprathreshold pulse amplitude and held constant between trials. Pulse frequency was set to 20 Hz and also did not vary between trials, with one exception during the phosphene shape mapping task that is noted below.

### Phosphene shape mapping task

Since phosphene shape and size differ between electrodes and across participants (Nanduri et al. 2008; Beyeler et al. 2019; Luo et al. 2016), which could influence the overall image produced by multi-electrode stimulation, we performed a systematic mapping experiment. We delivered current through individual electrodes and asked participants to draw the phosphene they observed on an electronic tablet. The participants were asked to focus on phosphene size and shape, rather than on brightness or the position in the visual field. We tested 25 electrodes per participant with 12 repetitions each. Pulse frequency was initially set to 20 Hz but then lowered to 6 Hz due to perceptual adaptation occurring too quickly. Stimulation pulse train length was 200 ms, thus 2-3 pulses were delivered per train.

To quantify the shapes of the elicited phosphenes, we used computational metrics frequently used in image processing. The built-in functions of “regionprops” in the MATLAB (MathWorks, Inc.; Natick, MA, USA) Image Processing toolbox were used to calculate area and elongation (also known as “eccentricity”). Area equaled the total number of non-zero pixels. An ellipse was fit to the drawn phosphene, and the elongation metric equaled the ratio of the distance between the foci of the fitted ellipse and its major axis length. Elongation was expressed as a fraction, where elongation=0 represented a perfect circle and elongation=1 represented a line. If two or more phosphenes were perceived in a single trial, only the first drawn phosphene was included when calculating elongation; it was assumed to be the most noticeable, and the computation cannot account for multiple phosphenes.

We ran several statistical analyses on the phosphene shape data using MATLAB. Significance levels in all statistical analyses were set to α=0.05 and Bonferroni corrections were applied. First, we ran one-way ANOVAs to compare area and elongation between electrodes within each participant. We also ran a linear regression to determine if there was a statistically significant relationship between mean phosphene area (across 12 trials per electrode) and detection threshold. Next, we performed computational analyses of phosphene shapes (Beyeler et al. 2019; Nanduri et al. 2008). We started by calculating the standard error of the mean (SEM) of area and elongation for each electrode. We then took 12 random samples, with replacement, from all 300 drawings (across all electrodes and trials) from each participant and calculated a resampled SEM across electrodes. We randomly selected one of the 25 SEMs that corresponded to single electrodes, and evaluated if it was less than the resampled SEM. We calculated this 1000 times and extracted the proportion of SEMs across electrodes that were larger than the SEMs within an electrode, which served as the p-value. This analysis was performed separately for each participant and for area and elongation.

### Identifying optimal sequential stimulation parameters

To determine which sequential stimulation paradigm would result in the clearest perception, we varied pulse train durations and gap intervals. The participants’ task was to answer if phosphenes were moving from right to left, left to right, bottom to top, or top to bottom. Three electrodes were stimulated to create each directional pattern. We set the pulse train duration of each electrode to 50, 100, 150, or 200 ms. We also tested gap intervals, or the amount of time between electrode stimulations, of 0, 50, and 100 ms. We performed three repetitions for each combination of train duration and gap interval, for a total of 48 trials per participant.

### Size discrimination task

We performed a two-alternative forced choice (2-AFC) task in which the participants were presented with two squares and had to choose which was larger. The squares were created by stimulating four electrodes within the array to form the four corners (Figure 1b). There were four different sizes of squares: each corner was separated by 1, 2, 3, or 4 electrodes. The comparison pairs alternated between trials. For participant S2, the electrodes were stimulated for 200 ms during the simultaneous stimulation paradigm; during the sequential stimulation paradigm, each electrode was stimulated for 200 ms with a 50 ms gap between electrodes. We initially set the same parameters for participant S1 as well, but perceptual adaptation occurred too quickly and train duration was subsequently set to 100 ms for both the sequential and simultaneous paradigms.

Sequential and simultaneous stimulation paradigms were each tested 36 times per participant. We attempted to select electrodes that elicited similar phosphene shapes and sizes, but some of the electrodes for the larger squares fell outside the bounds tested during the phosphene shape mapping experiment. Therefore, only a subset of the tested electrodes could be matched in size and shape. We ran a test of two proportions to evaluate if sequential stimulation would enable significantly better performance than simultaneous stimulation, a test of one proportion to evaluate if simultaneous stimulation performed better than chance, and another test of one proportion to evaluate if sequential stimulation performed better than chance.

### Simple shape discrimination task

In another 2-AFC task, participants were presented with an “arrow,” and had to respond if the arrow was pointing to the left or the right. The arrows were created by stimulating three electrodes within the array (Figure 1c). During the simultaneous stimulation paradigm, the electrodes were simultaneously stimulated for 200 ms. During the sequential stimulation paradigm, each electrode was stimulated for 200 ms. Since we did not find significant differences in performance between different gap intervals, we used a 0 ms gap between electrodes. Based on the results of the phosphene shape mapping task, we selected electrodes for participant S1 that elicited similar phosphene shapes and sizes. We did not do this for participant S2 because she was able to perform this experiment without it, and because the phosphene shapes elicited by different electrodes within her array had substantial variation that made it difficult to identify patterns. Sequential and simultaneous stimulation were each tested 20 times per participant. Three statistical tests, as described in the previous subsection, were performed for each participant to compare and contrast sequential vs. simultaneous stimulation.

### Complex shape discrimination task

We performed a 4-AFC task in which the participants were presented with a letter of the alphabet, and had to choose which letter it was. To minimize complexity, only a subset (O, V, K, Z) of the Sloan letters (Sloan, Rowland, and Altman 1952) were tested. Each letter was formed using 5-8 electrodes (Figure 1d). During the simultaneous stimulation paradigm, the electrodes were stimulated for 200 ms. During the sequential stimulation paradigm, each electrode was stimulated for 200 ms, with no gap (0 ms) between temporally adjacent electrodes. Each Sloan letter was tested 20 times with simultaneous stimulation and 20 times with sequential stimulation. Based on the results of the phosphene shape mapping task, we attempted to select electrodes that elicited similar phosphene shapes and sizes. Due to the limited ability of participants to successfully perform this task, the only statistical analysis we ran was a test of one proportion to evaluate if participant S2’s performance using sequential stimulation was significantly higher than chance.

## Results

### Phosphene shape mapping task

We found that different electrodes within each participant’s array elicited a variety of different phosphene shapes. The most commonly elicited shapes were circles, ovals, lines, and rectangles, though many shapes were irregular. Many of participant S1’s electrodes elicited similar phosphene shapes across trials (Figure 2a). For example, electrodes A04, A05, B08, and C06 usually elicited irregular ovals while electrodes D04, E04, E05, E07, and E08 typically elicited a thick line. The shapes across trials for participant S2 were far less reliable and did not follow any noticeable patterns (Figure 2b); 18/25 electrodes elicited at least three common geometric shapes, such as triangles, rectangles, ovals, trapezoids, and half circles.

**Figure 2:**
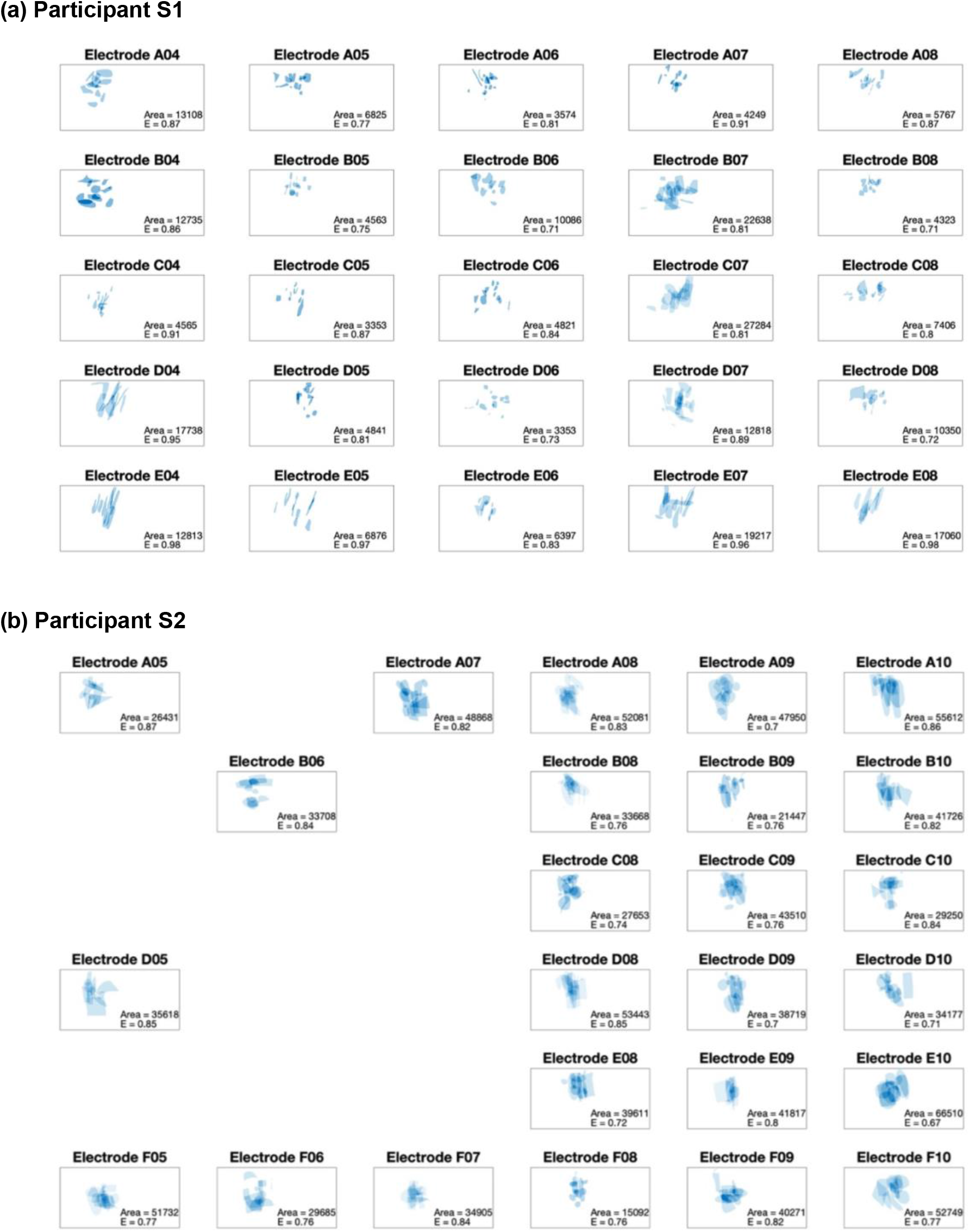
Phosphene shapes elicited by 25 electrodes, each tested 12x, for participants S1 **(a)** and S2 **(b)**. The phosphenes were drawn by each participant using a tablet with a touch screen. The resulting images are shown overlaid on top of each other, with darker blue regions indicating that 2+ phosphenes partially overlap. The mean phosphene area and elongation values across the 12 trials for each electrode is displayed in the bottom right corner of each subplot. The units of area are the number of pixels and elongation is in arbitrary units, with values closer to 1 signifying that the phosphene is shaped more similar to a line than to a circle.

It was most common for participants to report one phosphene per electrode (Figure 3a). Between the two participants, just seven electrodes produced 2+ phosphenes in at least 50% of trials. Phosphene area and elongation were significantly different between electrodes for participant S1 (p<0.001) but not S2 (Figure 3b,c). The variability in area and elongation was not more consistent within versus across electrodes for either participant (p>0.05). For participant S1 only, there was a statistically significant positive relationship between detection threshold and phosphene area (p<0.001, Figure 3d). Compared to participant S1, participant S2 tended to have larger phosphene areas and lower detection thresholds.

**Figure 3:**
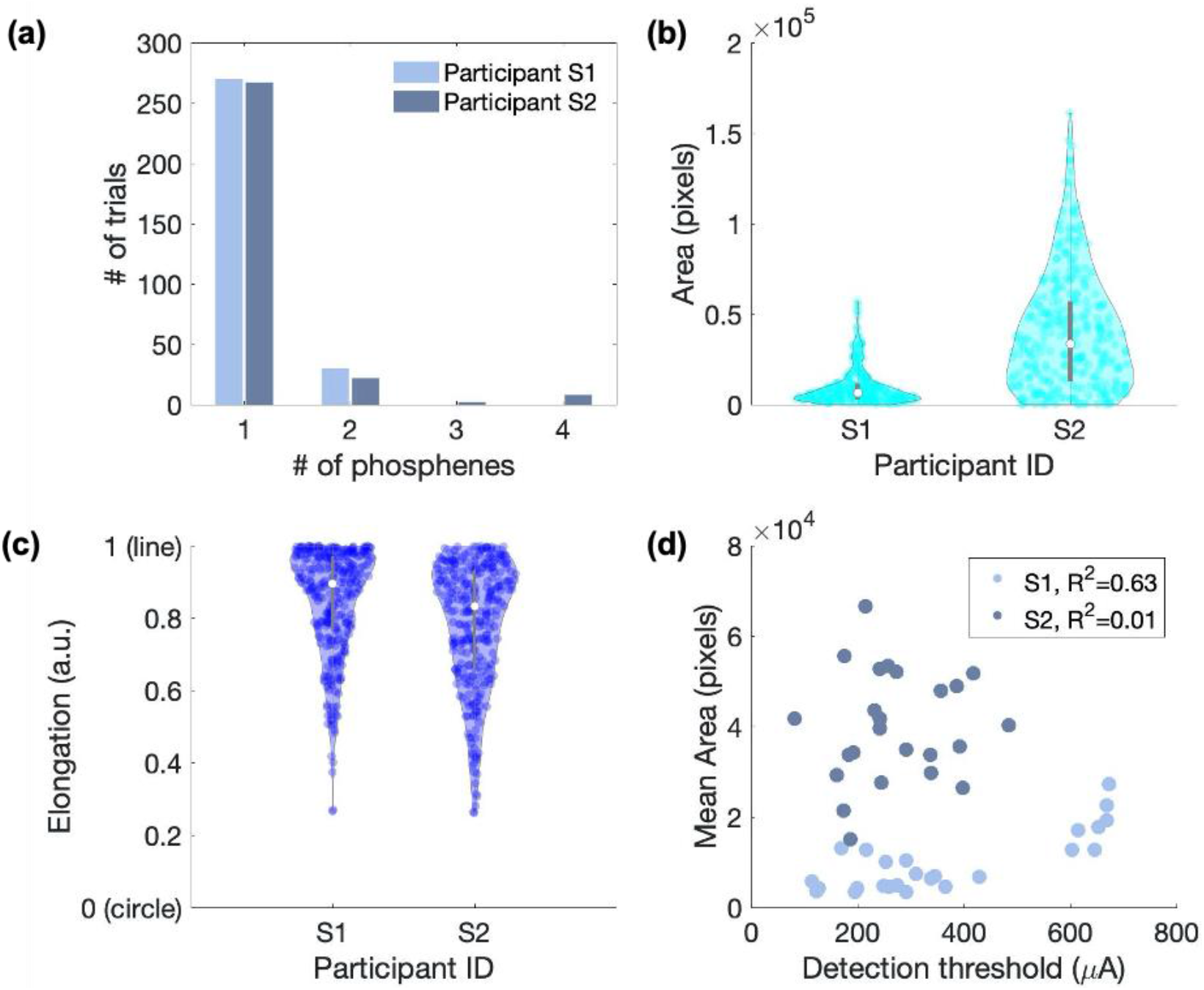
Computational metrics performed on phosphene shape data. **(a)** The number of phosphenes elicited by stimulating through a single epiretinal electrode. Collapsed across electrodes and trials for each participant. **(b)** The area of the phosphenes elicited by stimulating through a single electrode. Each point represents one trial (for a total of 12 trials * 25 electrodes = 300 trials per participant). **(c)** The elongation of the phosphenes elicited by stimulating through a single electrode. Elongation=0 represents a perfect circle and elongation=1 represents a line. **(d)** The mean phosphene area (across 12 trials) as a function of phosphene detection threshold. Each point represents one electrode. The R^2^ corresponding to a linear regression is displayed for each participant (S1 and S2).

### Sequential stimulation parameter evaluation

Classification accuracy of phosphene direction was highest (92% correct) when the train duration was set to 200 ms per electrode and lowest (71% correct) at 100 ms (Table 1). There were only negligible differences in performance as a result of changing the gap intervals between electrodes: classification accuracy had a minimum of 78% correct for 100 ms gaps and a maximum of 81% correct for 0 or 50 ms gaps. The best combination of stimulation parameters across both participants was a 200 ms train duration with either a 0 ms or 50 ms gap.

**Table 1:**
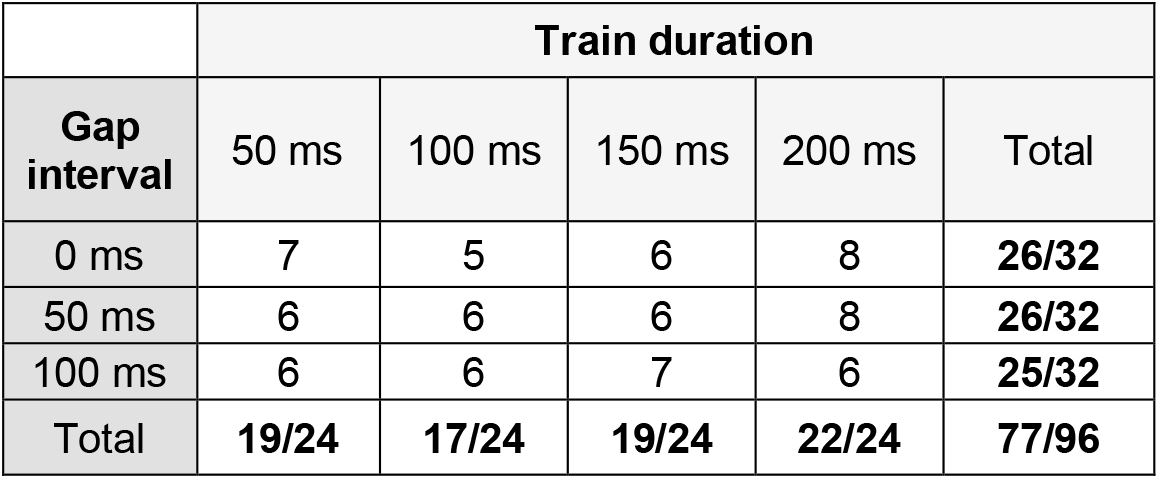
Sequential stimulation parameter tuning. The participants were presented with a pattern of phosphenes generated by stimulation through three electrodes. They were asked to respond if the phosphenes were moving left to right, right to left, top to bottom, or bottom to top. The stimulation pulse train duration and inter-electrode gap interval varied across the 48 trials per participant. Each element in the table depicts the number of trials that were answered correctly, collapsed across both participants.

### Size discrimination task

The participants’ performance on the square size discrimination task was better when using sequential versus simultaneous stimulation paradigms (Table 2), but the improvement was statistically significant only for participant S1 (p=0.01). Both participants achieved 27 out of 36 trials correct using sequential stimulation, which was significantly higher than 50% chance (p=0.001). When performing the task using simultaneous stimulation, participant S1 got 17/36 trials correct, which was not significantly higher than chance, and participant S2 got 25/36 trials correct, which was better than chance (p=0.001). When asked to draw what they perceived during sequential and simultaneous stimulation, the participants reported seeing four discrete phosphenes (forming the four “corners” of the square) during sequential stimulation only (Figure 4). When the four electrodes were simultaneously stimulated, the phosphenes merged and participants reported seeing only one or two large phosphenes.

**Table 2:** Performance values for both participants on the three discrimination tasks. During the size discrimination task, the human participants were asked to choose which square appeared larger. During the simple shape discrimination task, the individuals were asked to choose if an arrow was pointing to the left or to the right. During the complex shape discrimination task, the individuals were asked to report if a collection of phosphenes resembled the letter O, V, K, or Z. The number of correct trials for each condition and for each participant are given in the table.

|  | Participant S1 |  | Participant S2 |  |
| --- | --- | --- | --- | --- |
|  | *Simultaneous stimulation* | *Sequential stimulation* | *Simultaneous stimulation* | *Sequential stimulation* |
| **Size discrimination task** | 17/36 | 27/36 | 25/36 | 27/36 |
| **Simple shape discrimination task** | 4/20 | 19/20 | 8/20 | 19/20 |
| **Complex shape discrimination task** | Not possible | 34/80 | Not possible | Not possible |

**Figure 4:**
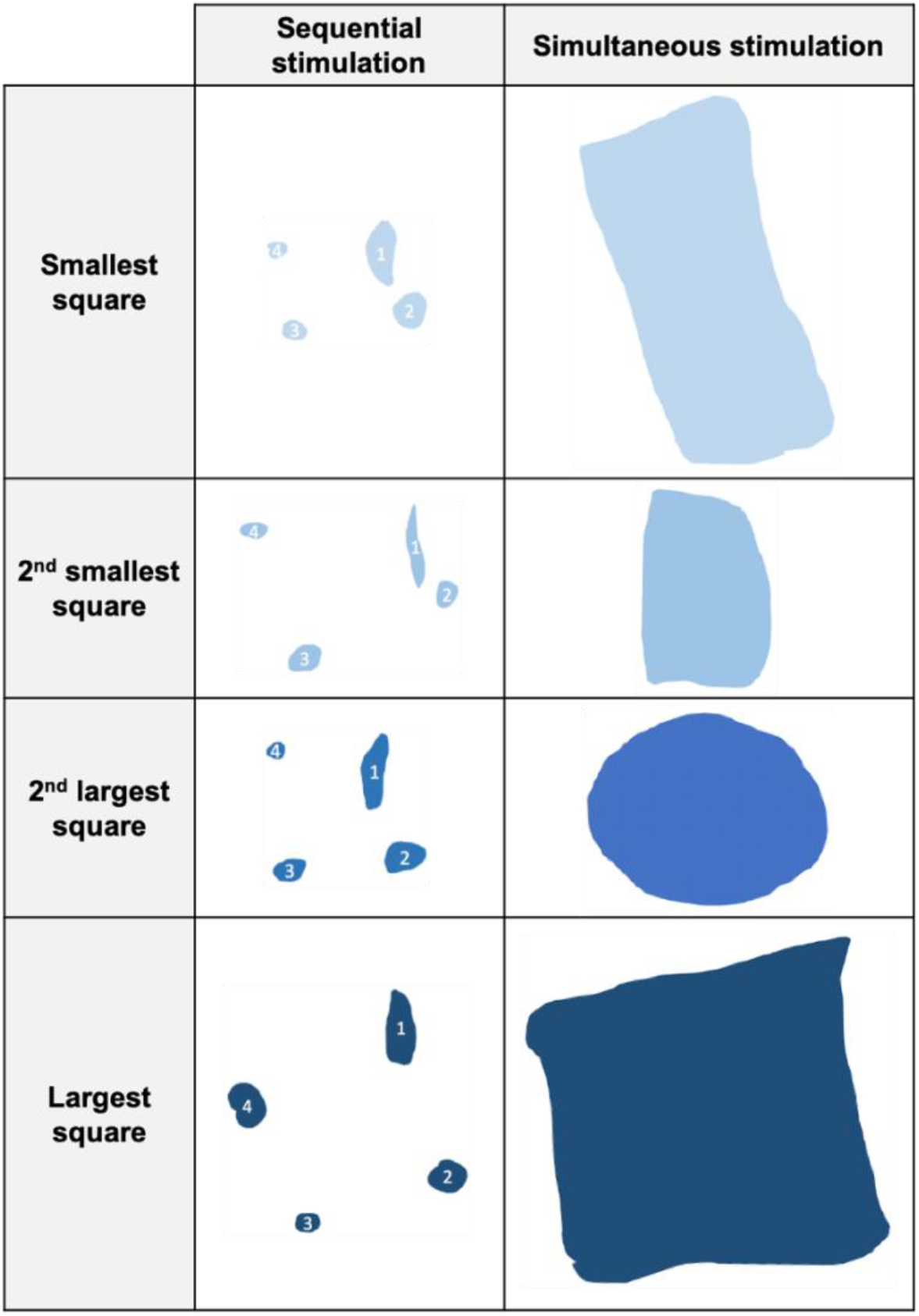
Participant S2’s drawings of what she perceived during the square size discrimination task. White numbers within phosphenes represent the order of presentation under the sequential stimulation paradigm. Though there were four electrodes stimulated during the simultaneous stimulation condition, the phosphenes merged into one shape that was sometimes difficult to interpret as a square.

### Simple shape discrimination task

The participants’ performance on the arrow discrimination task was significantly better when using sequential compared to simultaneous stimulation paradigms (p<0.001 for both participants, Table 2). Participant S1 could not originally perform this task until we selected electrodes that elicited similar phosphene shapes and sizes. Both participants correctly identified arrow direction in 19/20 trials when using sequential stimulation, which was significantly higher than 50% chance (p<0.001). When using simultaneous stimulation, participant S1 achieved 4/20 trials correct and participant S2 achieved 8/20 trials correct, neither of which were significantly better than chance (p>0.05). In fact, participant S1’s accuracy was far below chance; when presented with a right-pointing arrow using simultaneous stimulation, he reported that it was pointing to the left in 9/10 trials.

When the participants were asked to draw the shapes of the arrows, the three phosphenes merged together in an unpredictable manner during simultaneous stimulation (Figure 5). Though the participants saw three discrete phosphenes during the sequential stimulation paradigm, they reported seeing only 1-2 phosphenes when the three electrodes were simultaneously stimulated. When participant S1 was presented with a right-pointing arrow using simultaneous stimulation, two phosphenes merged into one larger phosphene on the right-hand side of his visual field. It is possible that participant S1 interpreted the larger phosphene as being the wider base of the arrow, and the smaller phosphene as the narrower tip of the arrow, and was inclined to report right-pointing arrows as left-pointing arrows during simultaneous stimulation trials.

**Figure 5:**
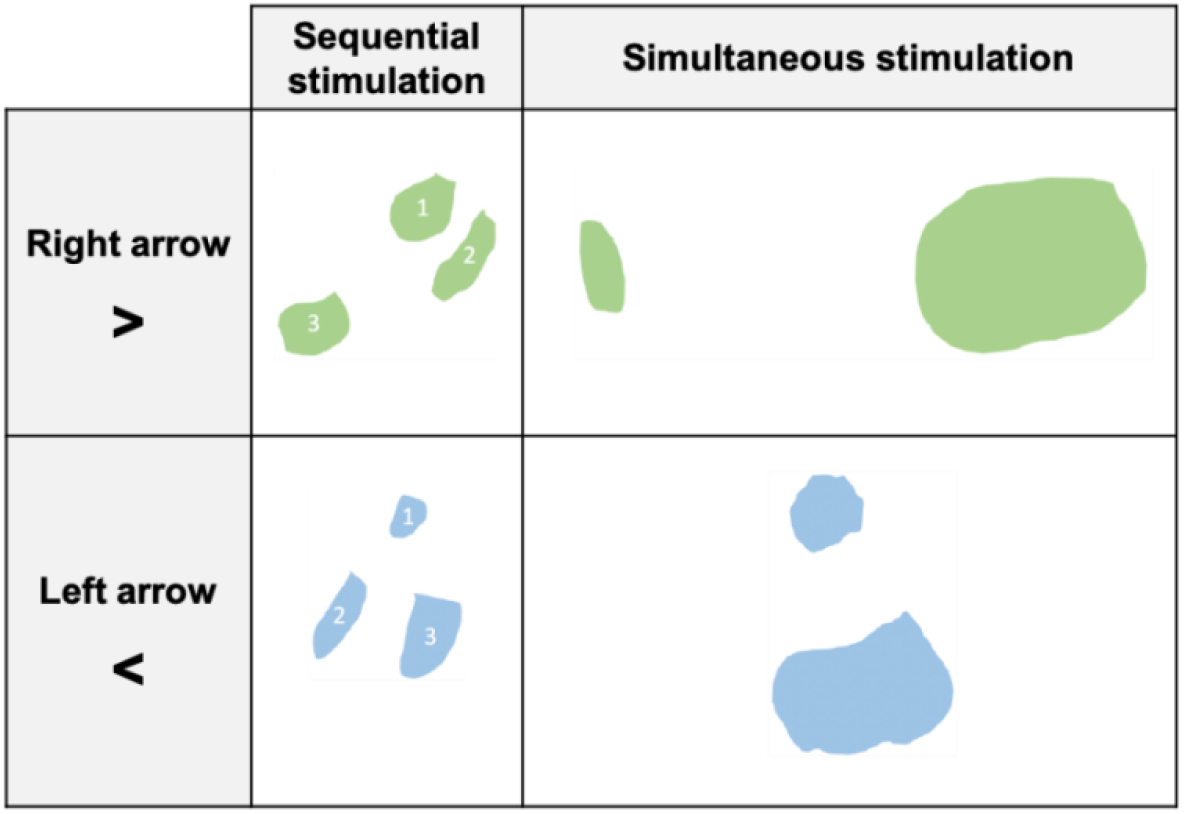
Participant S1’s drawings of what he perceived during the arrow discrimination task. Phosphenes shaded in green were what he drew when presented with a right-pointing arrow, and blue represents what he drew for a left-pointing arrow. White numbers within phosphenes represent the order of presentation under the sequential stimulation paradigm. Though there were three electrodes stimulated during the simultaneous stimulation condition, the phosphenes merged into two shapes that were difficult to interpret as an arrow.

### Complex shape discrimination task

Only participant S2 was successfully able to complete the alphabet letter discrimination task, and only when using the sequential stimulation paradigm. Participant S2 was able to correctly choose which alphabet letter was presented in 34/80 trials, which was significantly higher than the 25% chance (p<0.001), but not optimal for functional use. She had a 40% accuracy for identifying letter ‘K,’ 35% for ‘O,’ 50% for ‘V,’ and 45% for ‘Z.’ Anecdotally, the participant commented that letters ‘O’ and ‘Z’ were easiest for her to identify. Both participants commented that the task was difficult because individual phosphenes had different shapes and sizes, even though we attempted to select electrodes that elicited similar phosphenes.

## Discussion

The primary objective of this study was to evaluate if sequential stimulation of epiretinal electrodes would elicit clearer shapes than stimulating through multiple electrodes simultaneously. We performed phosphene shape mapping experiments and forced choice discrimination tasks to evaluate if the participants could discriminate square size, arrow direction, and letters of the alphabet. Contrary to our original hypothesis, sequential stimulation did not consistently outperform simultaneous stimulation in these discrimination tasks.

### Phosphene shapes varied between electrodes and participants

The appearance of phosphenes was highly variable across electrodes and participants, which is consistent with prior studies (Nanduri et al. 2008; Beyeler et al. 2019; Luo et al. 2016). Stimulation elicited irregular shapes in addition to circles, ovals, lines, and rectangles. Only a small cohort of electrodes elicited phosphenes that appeared as focal spots of light. It is likely that there are a variety of phosphene shapes because stimulating through an epiretinal electrode not only activates the cell bodies that are just beneath the electrode in the ganglion cell layer, but it also activates axon bundles that pass over the surface of the retina in the nerve fiber layer while en route to the optic nerve (Beyeler et al. 2019; Esler et al. 2018; Tahayori et al. 2014; Rizzo et al. 2003; Fried et al. 2009; Weitz et al. 2015). It may be possible to minimize this additional recruitment by increasing the pulse width of electrical stimulation (Granley and Beyeler 2021), though this has not yet been tested with human subjects. The inadvertent recruitment of axon bundles is one reason why it may be advantageous to target the visual cortex for visual neuroprostheses, because it has a different cellular makeup than the retina.

Unlike prior studies, we also noticed a substantial amount of variation between trials within a single electrode, especially for participant S2. Before starting this experiment, participant S2 mentioned that she did not necessarily think of phosphenes in terms of shapes, and speculated that she might find the task difficult. In general, small variations between trials were likely due to the fact that the participants were drawing without any visual feedback of traces made by their finger on the tablet screen. Phosphene shape variation across trials also could have been influenced by perceptual adaptation (Pérez Fornos et al. 2012) and gaze direction (Barry and Dagnelie 2016; Sabbah et al. 2014; Caspi et al. 2017; 2021). However, we took frequent breaks to minimize the likelihood of adaptation, and we asked the participants to maintain the same head and eye position between trials.

### Sequential stimulation performed better for simple discrimination tasks only

Sequential electrode stimulation outperformed simultaneous stimulation on the size discrimination task and the simple shape discrimination task, but not on the complex shape discrimination task. In all cases, sequential stimulation helped to prevent multiple phosphenes from merging into one uninterpretable phosphene. However, the utility of sequential stimulation decreased as the complexity of the presented shape increased. As additional electrodes were utilized to create a more complex shape, it was difficult to select electrodes that elicited phosphenes that were similar in size and shape. Participants reported that the differences in phosphenes made discrimination tasks, especially the alphabet task, very difficult to perform.

Additionally, even with sequential stimulation, the participants did not necessarily perceive the presented images as specific shapes. The overall image created by multiple electrodes was sometimes intuitive or explained and quickly understood, but most instances required practice trials, especially for participant S1. Moreover, sometimes the electrodes did not elicit phosphenes in the expected location in the visual field, even when accounting for the angle of the electrode array on the retina. Mislocalization can be attributed to eye movements that influence the location of the phosphene (Sabbah et al. 2014; Caspi et al. 2017; 2021), or inconsistencies in phosphene size and brightness. Mislocalization could easily turn a common geometric shape into an unrecognizable polygon, which often required us to select a different set of electrodes.

## Conclusion

In conclusion, sequential electrode stimulation outperformed simultaneous stimulation in simple discrimination tasks, but not in more complex discrimination tasks. Because our surroundings in daily life are visually complicated, sequential stimulation may be most beneficial for epiretinal prostheses in basic navigational applications rather than difficult tasks such as object identification. For example, sequential stimulation paradigms could be used to inform an Argus II user that they need to move in a specific direction to avoid an upcoming obstacle. It is possible that sequential stimulation will be more impactful for epiretinal prostheses once there is a way to carefully manipulate the phosphene shape elicited by each electrode. However, given the additional temporal requirements associated with sequential stimulation compared to simultaneous stimulation, performance would have to be incontrovertibly better in order for it to be adopted for functional applications.

## Data Availability

All data produced in the present study are available upon reasonable request to the authors.

## Acknowledgements

The authors would like to thank the research participants for their time, patience, and dedication. Research reported in this publication was supported by the National Eye Institute of the National Institutes of Health under Award Number R01EY029741. In addition, this work was supported with the resources and use of facilities at the Johns Hopkins Hospital.

## Disclaimer

The content is solely the responsibility of the authors and does not necessarily represent the official views of the National Institutes of Health.

